# Co-resistance between oral antibiotics for pyelonephritis and those for simple urinary tract infections – Applying an Escalation Antibiogram Model to local community data

**DOI:** 10.1101/2025.07.09.25331170

**Authors:** Philip Williams, Edward Barton, Ranjeet Bhamber, Léo Gorman, Andrew W Dowsey, Matthew B Avison

## Abstract

**Objectives:** The objective of this study was to apply an escalation antibiogram (EA) to community urine data to assess how presumptive resistance (treatment failure or recent microbiological samples) to first-line antibiotics for simple urinary tract infections (UTIs) affects resistance to antibiotics used to treat pyelonephritis. Furthermore, we examined how this varies with age or in instances of recurrent UTI.

**Methods:** We extracted susceptibility data from *Escherichia coli* isolates grown from urine samples sent from general practice from a 5-year period (2019–2023) in a region served by three NHS hospital trusts. All female patients over 18 years old were included giving a total of 130,514 isolates. We applied a Bayesian model to estimate antibiotic resistance rates for oral pyelonephritis antibiotics, when presuming resistance to each of the first-line antibiotics used to treat simple UTIs. The model estimates the probability of resistance with 95% credible intervals and was applied to a variety of patient groups based on age and history of recurrent UTIs. The uncertainty in these estimates increases for smaller patient groups.

**Results:** Resistance to first-line UTI antibiotics has a marked effect on the probability of resistance to oral antibiotics used to treat pyelonephritis. In particular amoxicillin-clavulanate should be avoided for pyelonephritis if resistance to pivmecillinam is presumed in UTI because resistant rates may exceed 50%. For patients with presumed resistance to nitrofurantoin or trimethoprim in UTI, the optimal pyelonephritis antibiotic depends on both age group and history of past infections. For example, for patients under 50 with recurrent UTIs, amoxicillin-clavulanate has the lowest estimated resistance rate, but for women over 65 with recurrent UTIs, ciprofloxacin is optimal for pyelonephritis, where there is presumed nitrofurantoin resistance in UTI, but cefalexin is superior if trimethoprim resistance is presumed.

**Conclusions:** EA analysis informed by our Bayesian model is a useful tool to support empiric antibiotic prescribing for pyelonephritis. It provides an estimate of local resistances rates and a comparison of antibiotic options with a measure of the uncertainty in the data.

## Introduction

Infections of the lower urinary tract (cystitis) in women are a common presentation to primary care (in the UK, particularly general practice [GP]) (1). A small proportion of these progress to infections of the upper urinary tract (pyelonephritis), which can lead to complications including renal scarring, bacteraemia and sepsis. While evidence suggests a significant proportion of cystitis would resolve without antibiotic treatment, clinical resolution is more likely when treated, and pyelonephritis is more common in cystitis not treated with antibiotics (2). Although not designed or powered to detect pyelonephritis, randomised controlled trials comparing cystitis antibiotic treatment with a placebo or non-steroidal anti-inflammatory drugs have demonstrated a lower rate of pyelonephritis in the antibiotic arm (2, 3, 4). An epidemiological study from Sweden of 752,289 women diagnosed with uncomplicated cystitis, showed the case rate for pyelonephritis following cystitis was 1.61% in patients not treated with antibiotics compared to 0.55% in those treated (5).

UK guidelines suggest first-line treatment for cystitis in adult women should be nitrofurantoin, trimethoprim, pivmecillinam or fosfomycin. Sending urine for microscopy, culture and sensitivities (MC&S) is only recommended in women under 65 for recurrent urinary tract infection (UTI), pregnancy, or if treatment failure or pyelonephritis is suspected (6).

If a patient initially treated for cystitis re-presents to their GP or to the emergency department with suspected pyelonephritis, they would be treated with one of the following oral antibiotics: cefalexin, ciprofloxacin, amoxicillin-clavulanate (co-amoxiclav), and, if in hospital, gentamicin would also be an option (7). Ciprofloxacin has been the mainstay of pyelonephritis treatment due to low resistance rates compared to other oral options and proven efficacy (8). Recently in the UK, concern about rare but severe side effects has led to a Medicines and Healthcare products Regulatory Agency warning that fluroquinolones should be avoided in mild or moderate infections, unless there is no suitable alternative (9). Hence while ciprofloxacin is still an available option, co-amoxiclav or cefalexin are likely to be preferred if they have comparable or acceptable rates of resistance. In pregnancy, ciprofloxacin should be avoided unless no safer option is available, due to arthropathy in animal studies (10). Co-amoxiclav is avoided near term due to a possible risk of necrotising enterocolitis in the neonate. (11)

While in some cases, treatment can be guided by recent urine susceptibility results, in many cases, pyelonephritis therapy would be empiric. Over a 4-year period from 2020 to 2023 our local emergency department at the Bristol Royal Infirmary treated 1,769 cases of pyelonephritis. Positive microbiology (from blood culture or a midstream urine sample) was only available in 30% of cases. This anecdotal account is supported by a published cohort study of 105 pyelonephritis cases from 2015, which found that 50% had positive microbiology (12).

Between 75% and 90% of pyelonephritis is caused by *E. coli*, and antibiotic guidelines for pyelonephritis are usually based on *E. coli* sensitivities (13). A number of studies have demonstrated that resistance to one antibiotic is associated with increased probability of resistance to other antibiotics, and three recent studies have investigated aspects of co-resistance in *E. coli* from UTI samples. Guy *et al* (14) explored the use of nitrofurantoin resistance as a marker for multi-drug resistance in samples from the UK. Kaye *et al* (15) described co-resistance between four antibiotic classes (nitrofurantoin, co-trimoxazole, fluoroquinolones and third generation cephalosporins) in the USA. Leroy *et al* (16) investigated co-resistance amongst oral antibiotics recommended for pyelonephritis in France concluding that an oral option is almost always available.

The term “escalation antibiogram” has been coined to describe an antibiogram generated to guide empirical therapy assuming resistance to other antibiotics (17). While the failure of initial cystitis treatment does not guarantee resistance to the initial antibiotic, the possibility of resistance should be considered when prescribing follow-up treatment. Accordingly, using local *E. coli* susceptibility data from GP urine samples, we applied a Bayesian model initially developed to guide empirical antibiotic escalations in suspected Gram-negative sepsis (18) to predict the resistance rate to antibiotics used locally for pyelonephritis treatment. We did this for scenarios where resistance to any one of our first-line cystitis antibiotics was assumed and where widespread susceptibility testing is performed (nitrofurantoin, trimethoprim and pivmecillinam).

Our model provides an expected value (posterior mean) with 95% credible interval to illustrate uncertainty including that from the size of the patient subgroup and can estimate probability of inferiority between two antibiotic options. This model can be applied to specific patient groups where resistance rates and underlying microbiology may differ from the general population. While the model is applicable to any patient group, in this paper we have used data from female patients, as this group has the highest burden of cystitis and pyelonephritis.

## Methods

Data were collected from a region served by three NHS trusts covering four acute hospitals in the Southwest of England (Royal United Hospital Bath NHS Foundation Trust, University Hospitals Bristol & Weston NHS Foundation Trust, and North Bristol NHS Trust) which share a single laboratory information management system (LIMS) (Winpath Enterprise 7.23, Clinisys). All *E. coli* grown from urine samples sent from GPs over a 5-year period 2019 to 2023, inclusive, in this region were included in the cystitis (nitrofurantoin, trimethoprim, and pivmecillinam) and pyelonephritis (co-amoxiclav, cefalexin, ciprofloxacin and gentamicin) antibiotic datasets. For each isolate, the antibiotic susceptibility profile was determined using European Committee on Antimicrobial Susceptibility Testing (EUCAST) disc testing methodologies. Results were expressed as “Susceptible” or “Resistant” with intermediate results classed as resistant. We extracted these susceptibility data on female patients over 18 years old giving a total of 130,514 isolates. Recurrent UTI was defined as 3 or more urine samples in a 12 month period. Data from 2018 were used to identify this patient subgroups in the 2019 data.

Predicted antibiotic resistance rates for pyelonephritis antibiotics with 95% credible intervals were calculated for the end of December 2023 based the previous 5 years data for each patient sub-group.

We use a Bayesian spline model with a Bernoulli likelihood and logit link function to model AST resistance rates over time, which is fitted by Hamiltonian Monto Carlo - please see our previous paper for more details (18). Resistance rates between two groups or time points were compared by subtracting one set of posterior predictive curves from another, allowing us to calculate the posterior probability of an increase (PPI) or decrease (PPD) in resistance over time, or to calculate posterior probability of inferiority (PPInf) or superiority (PPSup) between two antibiotic options.

## Results and Discussion

### Subgroup Selection

For female patients over 18 years within our region, resistance rates for *E. coli*, from GP-submitted urine samples for ciprofloxacin, cefalexin, co-amoxiclav and gentamicin were sufficiently low that any of these three antibiotics could be used empirically to treat pyelonephritis. See Table 1.

**Table 1.** Posterior mean resistance estimates for urinary E. coli with 95% credible interval for pyelonephritis antibiotics, for all female (All F) patients, from GP samples.

| Age Group | UTI samples | Number | Cefalexin | Ciprofloxacin | Coamoxiclav | Gentamicin |
| --- | --- | --- | --- | --- | --- | --- |
| 18 to 50 | All | 30,572 | 0.08 (0.07 - 0.09) | 0.06 (0.05 - 0.07) | 0.08 (0.07 - 0.10) | 0.04 (0.03 - 0.05) |
|  | First | 23,590 | 0.07 (0.06 - 0.08) | 0.05 (0.04 - 0.06) | 0.08 (0.07 - 0.09) | 0.04 (0.03 - 0.05) |
|  | Two Plus | 2,552 | 0.12 (0.08 - 0.16) | 0.10 (0.07 - 0.14) | 0.09 (0.06 - 0.13) | 0.07 (0.03 - 0.10) |
|  | Pregnant | 2,557 | 0.09 (0.05 - 0.13) | 0.04 (0.07 - 0.12) | 0.11 (0.07 - 0.17) | 0.04 (0.02 - 0.07) |
| 51 to 65 | All | 22,994 | 0.07 (0.06 - 0.08) | 0.06 (0.05 - 0.07) | 0.09 (0.07 - 0.10) | 0.05 (0.04 - 0.06) |
|  | First | 15,084 | 0.07 (0.05 - 0.08) | 0.05 (0.04 - 0.06) | 0.08 (0.10 - 0.07) | 0.05 (0.04 - 0.06) |
|  | Two Plus | 3,647 | 0.09 (0.06 - 0.15) | 0.09 (0.06 - 0.12) | 0.11 (0.08 - 0.14) | 0.08 (0.05 - 0.12) |
| Over 65 | All | 66,567 | 0.07 (0.06 - 0.08) | 0.07 (0.06 - 0.08) | 0.09 (0.08 - 0.10) | 0.05 (0.05 - 0.06) |
|  | First | 34,455 | 0.06 (0.05 - 0.07) | 0.06 (0.05 - 0.07) | 0.07 (0.06 - 0.09) | 0.04 (0.03 - 0.05) |
|  | Two Plus | 17,288 | 0.09 (0.08 - 0.11) | 0.09 (0.08 - 0.10) | 0.12 (0.10 - 0.13) | 0.07 (0.06 - 0.09) |

Using our escalation antibiogram approach (18) we investigated how resistance to first-line cystitis antibiotics (nitrofurantoin, trimethoprim and pivmecillinam) among urinary *E. coli* alters the expected resistance rates of pyelonephritis antibiotics for all patients and in a range of clinically important sub-groups. Our methodology can be applied to any subgroup of clinical interest that can be identified within the available pathology meta-data. The smaller the group of interest, the greater the uncertainty in the results, which is reflected in the wider 95% credible intervals, but the data may more accurately reflect the microbiology of that group and its antibiotic exposures.

We investigated how the optimal pyelonephritis treatment options vary with age group and number of positive urine samples sent per patient in the previous 12 months, which we use as a proxy for prior antibiotic exposure. Pyelonephritis is most common in women between 18 and 29 years old and rates increase again from the age of 50 onwards (19). For women over 65 years, UTIs become increasing common with a rate of infection twice that of the group as a whole (20). This is associated with higher rates of resistance and, in addition to this, urine samples are routinely sent in this age group, rather than only being sent for complex or recurrent infections. To reflect these differences in epidemiology and sample collection we have considered three age groups in our research: 18-50, 51–65 and over 65 years. In addition to looking at all urine samples for each age group, we also looked at the first sample sent for each patient in a 12-month period (i.e. no prior samples) and at patients with recurrent UTI, defined as 3 or more urine samples being sent with a 12-month period (i.e. 2 or more prior samples). Note that the first sample sent for each patient in the under 65 age group may not represent their first UTI, so our data may overestimate the resistance expected for a patient’s actual first presentation.

The results for all samples and the selected sub-groups are seen in Table 1 to 4. When comparing antibiotic options for a large data set, a small difference in resistance rate can be reproduceable but not clinically significant. For example, in *E. coli* positive urine samples from women over 65 (n=66567), where there is resistance to trimethoprim, the model estimates that 15.8% (95% CI 14.1% to 17.6%) will be co-resistant to ciprofloxacin and 17.0% (95% CI 14.2% to 19.4%) co-resistant to co-amoxiclav. We can calculate it is 90% likely that resistance in ciprofloxacin is lower in this trimethoprim-resistant population, but this difference is of little clinical significance. See Figure 1 and Table 4.

**Figure 1.**
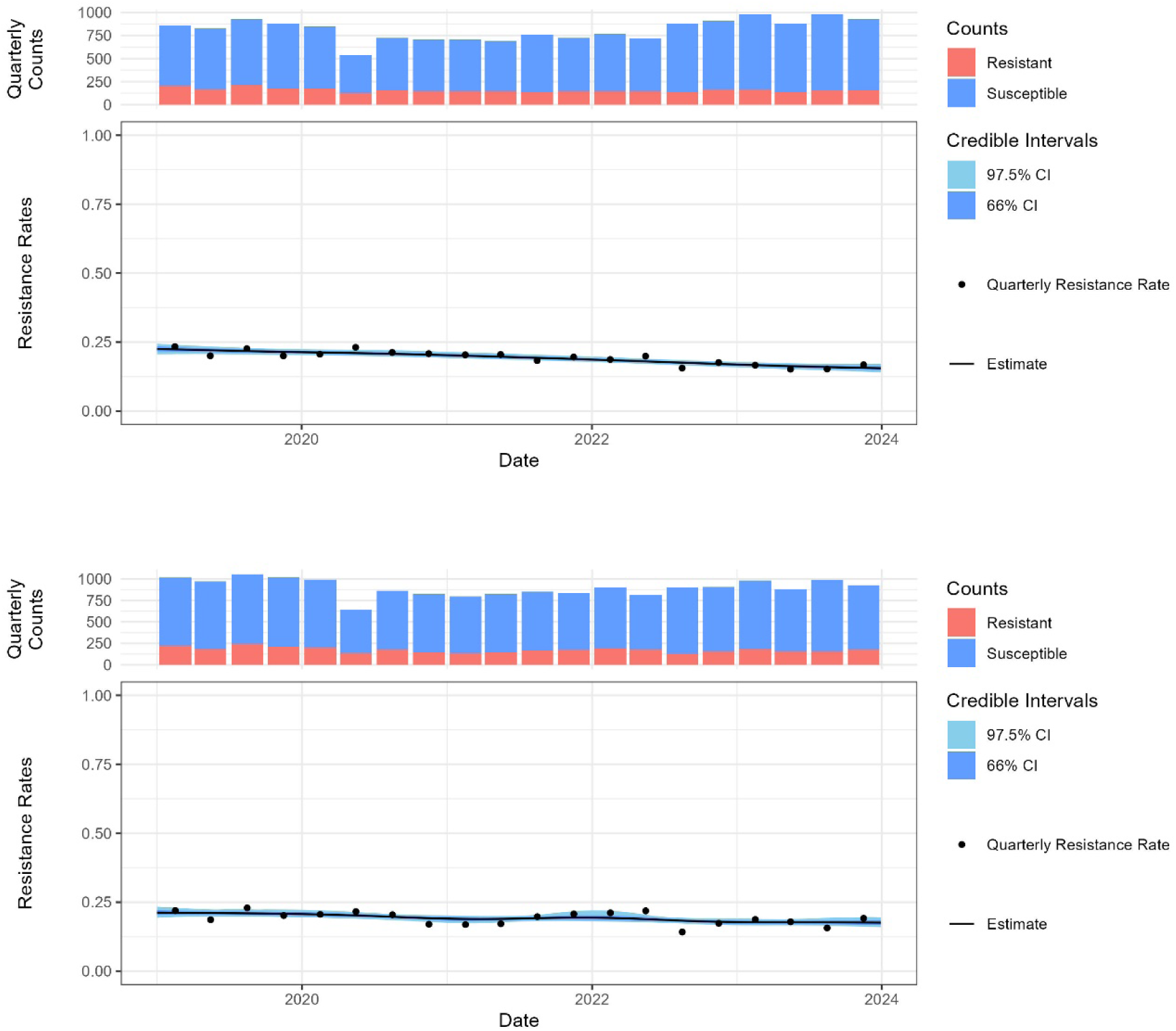
Percent Resistance to ciprofloxacin (Top) and co-amoxiclav (Bottom) assuming trimethoprim resistance, in all samples from women over 65 years of age. The black line shows the posterior probability of resistance and the shading the 66% and 97.5% credible intervals.

For small patient groups, where there is greater uncertainty in the data, a direct comparison between two antibiotic options may demonstrate similar probability of success despite an apparent difference in the posterior probability of resistance. For example, in *E. coli* positive urine samples from pregnant women (n=2499) with resistance to trimethoprim, 15.4% (95% CI 6.4% to 28.7%) are predicted to be co-resistant to ciprofloxacin and 20.5% (95% CI 11.7% to 32.5%) to cefalexin. In this case despite a difference of 5 percentage points in predicted resistant rates, the credible intervals overlap to the extent that there is a probability of 23.7% that resistance is actually lower for cefalexin. See Figure 2.

**Figure 2.**
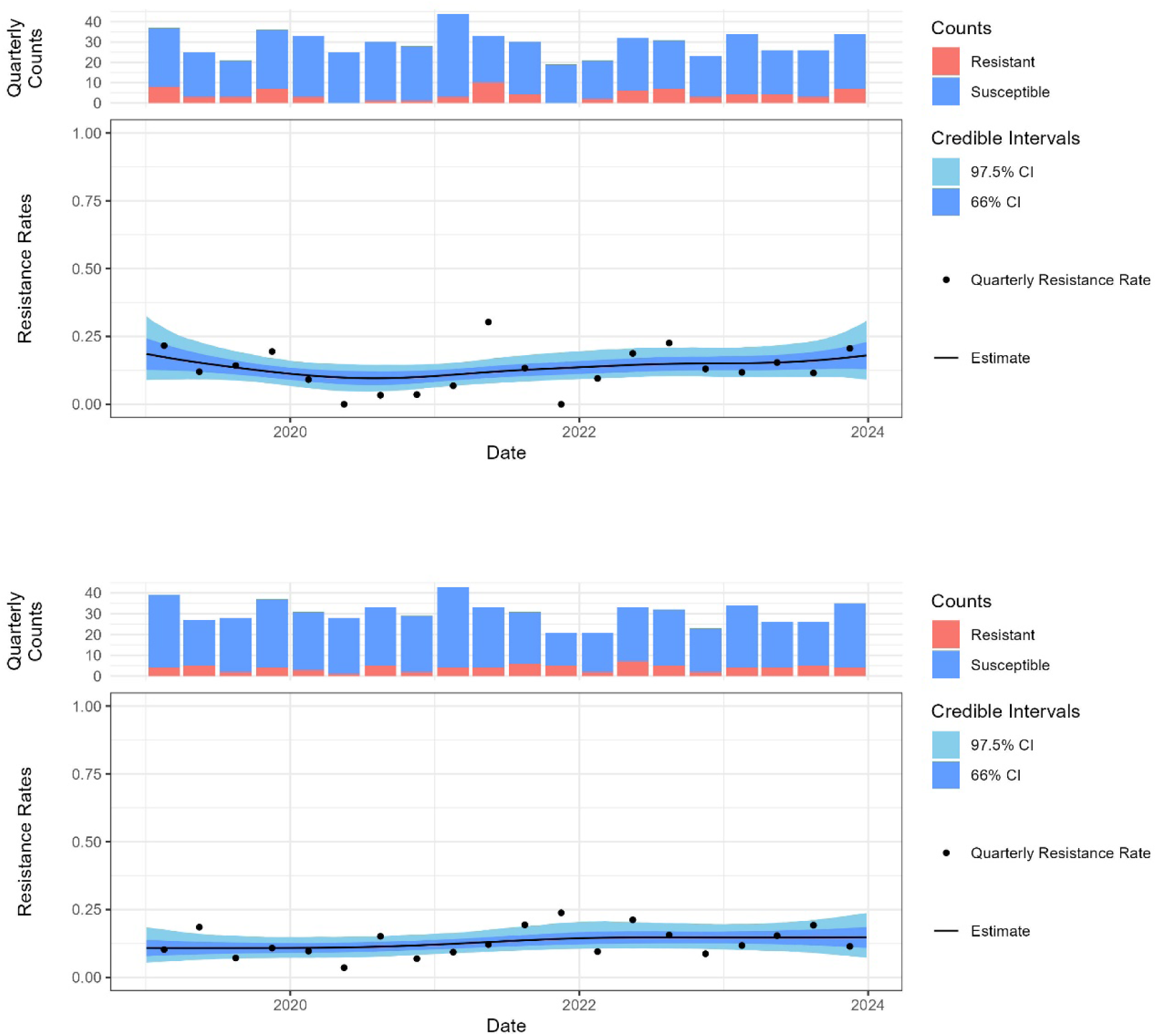
Percent Resistance to ciprofloxacin (Top) and co-amoxiclav (Bottom) assuming trimethoprim resistance, in pregnant women. The black line shows the posterior probability of resistance and the shading the 66% and 97.5% credible intervals.

We will now discuss the consequence of resistance to each of the three cystitis antibiotics in turn, assessed using our Bayesian approach, on predicted resistance to pyelonephritis antibiotics. These analyses are informed by microbiology data at population level. Their application to the treatment of individual patients would be guided by likely resistance to a first-line cystitis antibiotic, indicated by treatment failure, or by the observation of resistance in a recent urine sample.

### Pivmecillinam

In patients where pivmecillinam resistance is likely, co-amoxiclav should be avoided as expected resistance rates are between 50% and 60%, see Table 2. Hyper-production of TEM-1 beta-lactamase in *E. coli* has been reported to confer resistance to both co-amoxiclav and pivmecillinam (21), so this may explain the high rate of co-resistance observed here. See Figure 3.

**Figure 3.**
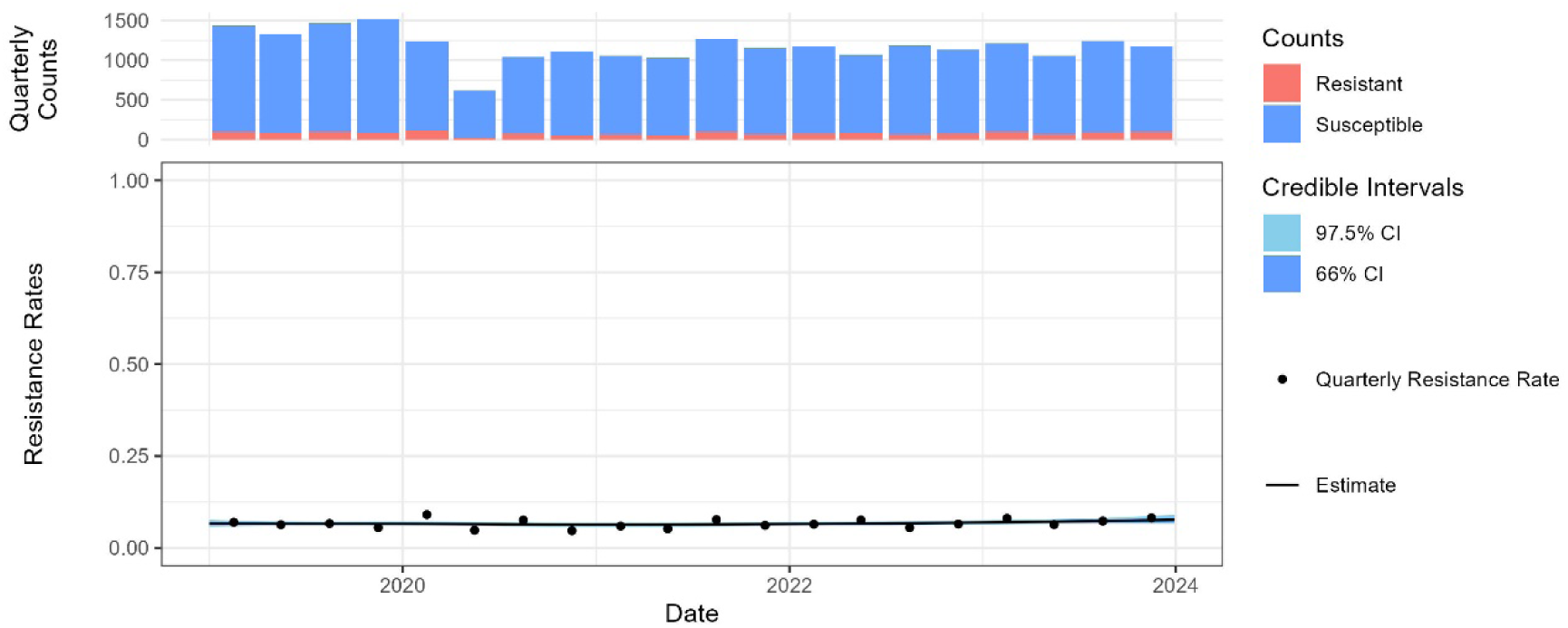
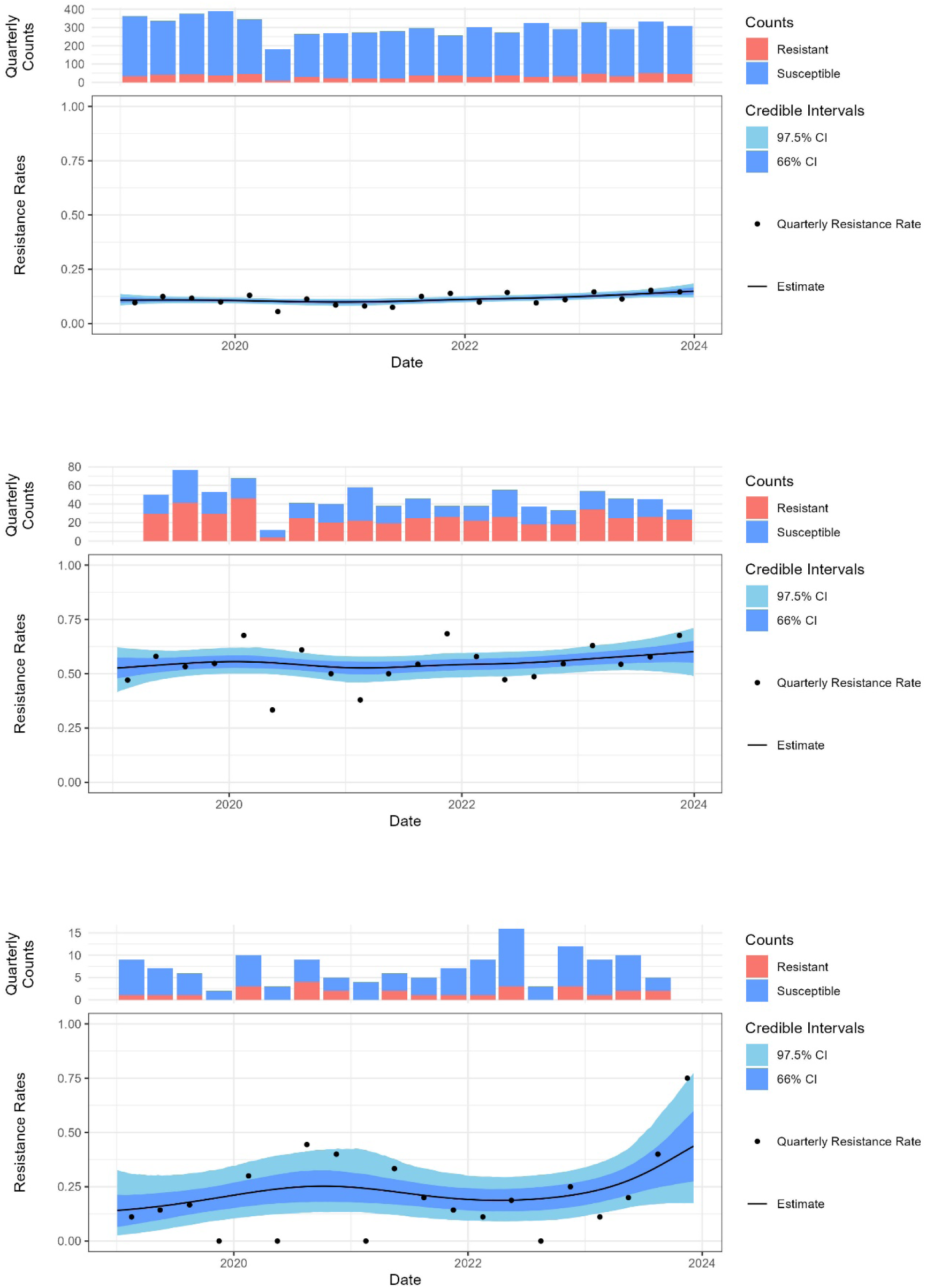
Predicted resistance to co-amoxiclav in women 18 to 50 years with no prior UTIs in the previous 12 months. (a) overall resistance, (b) assuming resistance to trimethoprim, (c) assuming resistance to pivmecillinam, (d) assuming resistance to nitrofurantoin. The black line shows the posterior probability of resistance and the shading the 66% and 97.5% credible intervals.

**Table 2.** Posterior mean resistance estimates for urinary E. coli with 95% credible interval for pyelonephritis antibiotics, given resistance to pivmecillinam, varying by age and number of UTIs the previous 12 months.

| Age Group | UTI samples | Number | Cefalexin | Ciprofloxacin | Coamoxiclav | Gentamicin |
| --- | --- | --- | --- | --- | --- | --- |
| 18 to 50 | All | 30,572 | 0.09 (0.05 - 0.17) | 0.06 (0.02 - 0.11) | 0.59 (0.48 - 0.72) | 0.12 (0.07 - 0.18) |
|  | First | 23,590 | 0.09 (0.04 - 0.19) | 0.04 (0.01 - 0.09) | 0.60 (0.48 - 0.72) | 0.11 (0.06 - 0.17) |
|  | Two Plus | 2,552 | 0.17 (0.04 - 0.42) | 0.15 (0.01 - 0.39) | 0.50 (0.26 - 0.77) | 0.23 (0.06 - 0.48) |
| 19 to 50 | Pregnant | 2,557 | 0.10 (0.01 - 0.29) | 0.05 (0.00 - 0.19) | 0.62 (0.37 - 0.86) | 0.16 (0.02 - 0.38) |
| 51 to 65 | All | 22,994 | 0.09 (0.05 - 0.15) | 0.08 (0.04 - 0.13) | 0.57 (0.48 - 0.66) | 0.12 (0.07 - 0.18) |
|  | First | 15,084 | 0.08 (0.03 - 0.14) | 0.08 (0.03 - 0.14) | 0.53 (0.41 - 0.64) | 0.11 (0.05 - 0.19) |
|  | Two Plus | 3,647 | 0.09 (0.02 - 0.23) | 0.08 (0.01 - 0.20) | 0.48 (0.32 - 0.66) | 0.16 (0.04 - 0.40) |
| Over 65 | All | 66,567 | 0.10 (0.07 - 0.13) | 0.14 (0.10 - 0.17) | 0.51 (0.42 - 0.59) | 0.12 (0.09 - 0.17) |
|  | First | 34,455 | 0.09 (0.05 - 0.14) | 0.12 (0.08 - 0.18) | 0.49 (0.39 - 0.57) | 0.11 (0.07 - 0.16) |
|  | Two Plus | 17,288 | 0.13 (0.09 - 0.19) | 0.15 (0.09 - 0.22) | 0.59 (0.48 - 0.70) | 0.15 (0.24 - 0.09) |

The expected resistance rates to ciprofloxacin and cefalexin were acceptable in all patient groups when resistance to pivmecillinam was present. Assuming no contraindications, cefalexin would generally be preferred given its favourable side effects profile. The highest rates of expected resistance were in patients 18 to 50 years with recurrent UTIs. Here cefalexin resistance was 16.7% (95% CI 3.5% to 42%) and ciprofloxacin resistance was 15.1% (95% CI 1.2% to 39%). This difference is unlikely to be clinically significant. See Table 2.

Given resistance to pivmecillinam, we found the expected resistance rate in pregnant women was 10% (95% CI 1.3% to 29%) for cefalexin compared to 5.4% (95% CI 0.4% to 18.6%) for ciprofloxacin. Cefalexin would still be preferred in this patient group. See Table 2.

Figure 4 shows how the model can be used to graphically display the probability of resistance to the four pyelonephritis antibiotics being considered here, and to calculate the probability of greater resistance to 3 of these compared with that with the lowest rate of resistance (ciprofloxacin). This example looks at women 18 to 50 years of age assuming pivmecillinam resistance, and with no prior UTIs or with recurrent UTIs. We can see that for those with no prior UTIs, ciprofloxacin has the lowest expected resistance rate, but that cefalexin is an acceptable option, while for those with recurrent UTIs, ciprofloxacin and cefalexin have a similar but higher rate of expected resistance.

**Figure 4.**
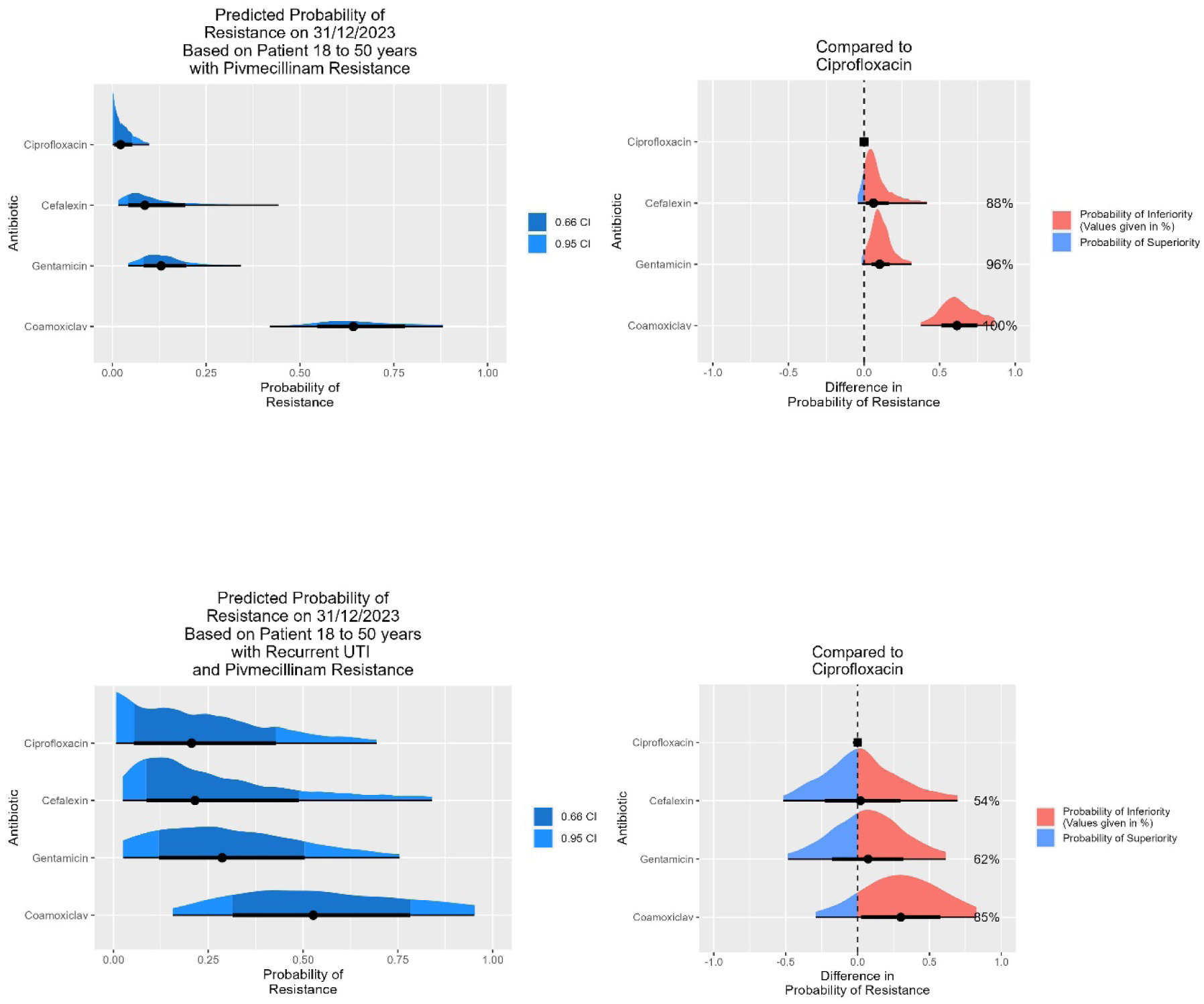
Predicted resistance to four pyelonephritis antibiotics assuming resistance to pivmecillinam with credible intervals, with a comparison of each antibiotic compared to ciprofloxacin. Female patients 18 to 50 age group with no prior UTIs (Top) and with recurrent UTIs (Bottom)

### Nitrofurantoin

Nitrofurantoin resistance is rare in urinary *E. coli* from women under 50 in our data, with an overall resistance rate of 1% (95% CI 0.5% to 1.5%), rising to 5.5% (95% CI 3.2% to 8.6%) in those with recurrent UTIs. This small resistant population results in higher uncertainty (thus wider 95% CI) in expected resistance to pyelonephritis antibiotics. See Figure 3.

Resistance rates, in this age group, were expected to be lowest for co-amoxiclav when resistance to nitrofurantoin was present. For a first episode of UTI all three oral options (co-amoxiclav, cefalexin and ciprofloxacin) have a similar expected resistance rate. See Table 3.

**Table 3.** Posterior mean resistance estimates for urinary E. coli with 95% credible interval for pyelonephritis antibiotics, given resistance to nitrofurantoin, varying by age and number of UTIs the previous 12 months.

| Age Group | UTI samples | Number | Cefalexin | Ciprofloxacin | Coamoxiclav | Gentamicin |
| --- | --- | --- | --- | --- | --- | --- |
| 18 to 50 | All | 30,572 | 0.20 (0.09 - 0.36) | 0.35 (0.20 - 0.52) | 0.16 (0.05 - 0.31) | 0.14 (0.04 - 0.29) |
|  | First | 23,590 | 0.26 (0.09 - 0.54) | 0.26 (0.07 - 0.53) | 0.24 (0.07 - 0.56) | 0.05 (0.00 - 0.20) |
|  | Two Plus | 2,552 | 0.21 (0.03 - 0.45) | 0.43 (0.10 - 0.75) | 0.11 (0.01 - 0.33) | 0.33 (0.08 - 0.63) |
|  | Pregnant | 2,557 | 0.49 (0.10 - 0.84) | 0.67 (0.25 - 0.98) | 0.47 (0.14 - 0.85) | 0.80 (0.39 - 0.99) |
| 51 to 65 | All | 22,994 | 0.16 (0.06 - 0.28) | 0.08 (0.00 - 0.21) | 0.19 (0.08 - 0.33) | 0.06 (0.02 - 0.17) |
|  | First | 15,084 | 0.28 (0.11 - 0.53) | 0.15 (0.02 - 0.39) | 0.12 (0.01 - 0.34) | 0.05 (0.00 - 0.18) |
|  | Two Plus | 3,647 | 0.09 (0.01 - 0.27) | 0.07 (0.00 - 0.39) | 0.29 (0.09 - 0.68) | 0.15 (0.01 - 0.64) |
| Over 65 | All | 66,567 | 0.29 (0.20 - 0.40) | 0.17 (0.11 - 0.25) | 0.23 (0.16 - 0.33) | 0.14 (0.08 - 0.25) |
|  | First | 34,455 | 0.11 (0.03 - 0.21) | 0.10 (0.04 - 0.18) | 0.16 (0.07 - 0.28) | 0.13 (0.05 - 0.26) |
|  | Two Plus | 17,288 | 0.39 (0.26 - 0.55) | 0.23 (0.13 - 0.37) | 0.28 (0.17 - 0.42) | 0.12 (0.05 - 0.22) |

In pregnant women where nitrofurantoin resistance was suspected, all pyelonephritis treatment options (including intravenous gentamicin) had a high rate of expected resistance with a wide 95% CI. For example, cefalexin resistance rate was estimated to be 49% but with 95% CI 10% to 84%. While cefalexin is most likely to be the best option (lower risk in pregnancy), the high resistance rate and high level of uncertainty suggests careful monitoring of the patient’s clinical status and a low threshold for hospital admission would be appropriate. Note nitrofurantoin should be avoided near term but may be used at other stages of pregnancy.

Given resistance to nitrofurantoin, in the older patient groups, our data suggest co-amoxiclav and ciprofloxacin would be favoured over cefalexin. See Table 3.

### Trimethoprim

In our region, trimethoprim resistance was common in urinary *E. coli*, varying from 26.4% (95% CI 24 % to 28.7%) in women under 50 years with a first UTI to 38% (95% CI 33.5 % to 42.9%) for women over 65 years with recurrent UTIs. This larger population positive for resistance means a narrow 95% CI for expected resistance to pyelonephritis treatments. See Figure 3.

Expected resistance rates were lower for ciprofloxacin than for cefalexin or co-amoxiclav for patients under 65 years where trimethoprim resistance is likely. Due to its potential side effects, co-amoxiclav or cefalexin are likely to be favoured when resistance rates are low enough for these agents to be viable options.

Cefalexin would be favoured in pregnancy despite a higher expected resistance rate of 20.5% (95% CI 11% to 32%) than ciprofloxacin at 15.4% (95% CI 6.4% to 28.7%), due to high level of uncertainty (there a probability of 23.7% that resistance to cefalexin is actually lower, see Figure 2) and possible risk of adverse fetal outcomes. Trimethoprim is contraindicated in the first trimester but can be used later in pregnancy.

For patients over 50 years old with trimethoprim-resistant organisms cefalexin had lower expected resistance rates than ciprofloxacin, with co-amoxiclav having the highest rates of resistance. See Table 4.

**Table 4.**
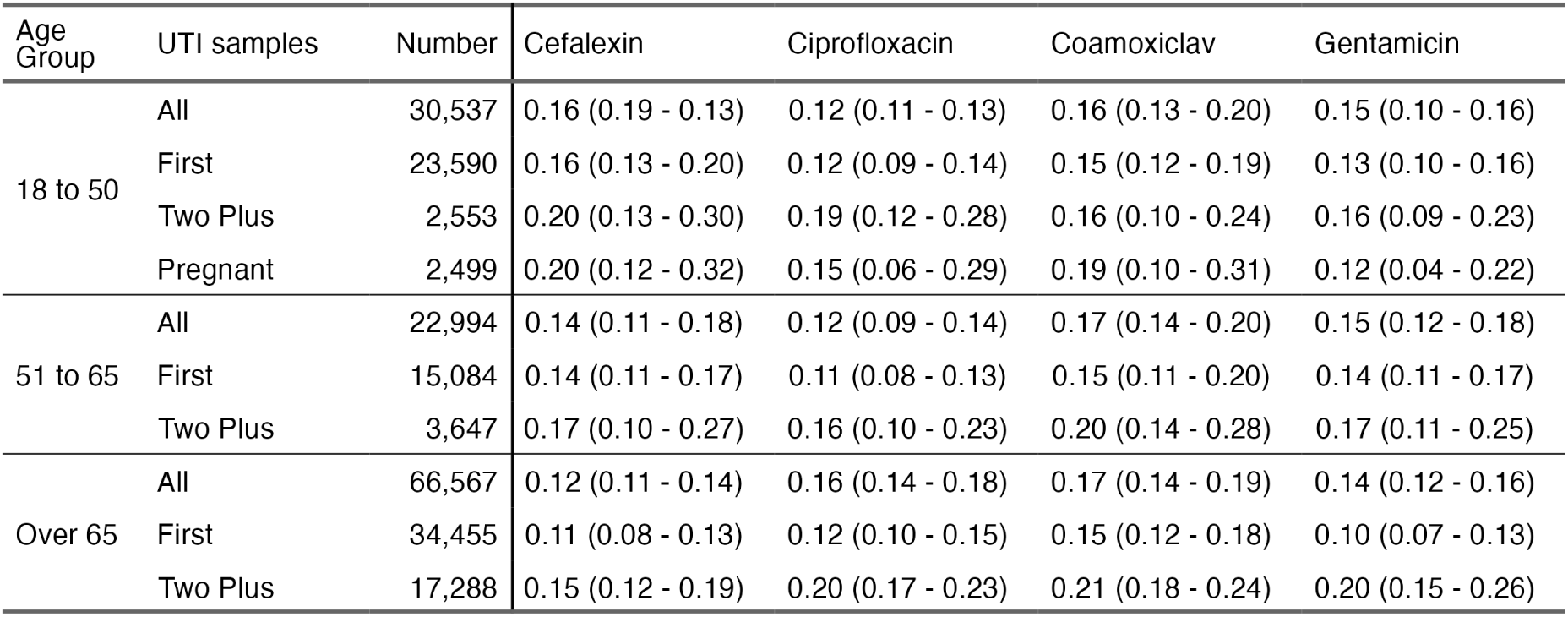
Posterior mean resistance estimates for urinary E. coli with 95% credible interval for pyelonephritis antibiotics, given resistance to trimethoprim, varying by age and number of UTIs the previous 12 months.

| Age Group | UTI samples | Number | Cefalexin | Ciprofloxacin | Coamoxiclav | Gentamicin |
| --- | --- | --- | --- | --- | --- | --- |
| 18 to 50 | All | 30,537 | 0.16 (0.19 - 0.13) | 0.12 (0.11 - 0.13) | 0.16 (0.13 - 0.20) | 0.15 (0.10 - 0.16) |
|  | First | 23,590 | 0.16 (0.13 - 0.20) | 0.12 (0.09 - 0.14) | 0.15 (0.12 - 0.19) | 0.13 (0.10 - 0.16) |
|  | Two Plus | 2,553 | 0.20 (0.13 - 0.30) | 0.19 (0.12 - 0.28) | 0.16 (0.10 - 0.24) | 0.16 (0.09 - 0.23) |
|  | Pregnant | 2,499 | 0.20 (0.12 - 0.32) | 0.15 (0.06 - 0.29) | 0.19 (0.10 - 0.31) | 0.12 (0.04 - 0.22) |
| 51 to 65 | All | 22,994 | 0.14 (0.11 - 0.18) | 0.12 (0.09 - 0.14) | 0.17 (0.14 - 0.20) | 0.15 (0.12 - 0.18) |
|  | First | 15,084 | 0.14 (0.11 - 0.17) | 0.11 (0.08 - 0.13) | 0.15 (0.11 - 0.20) | 0.14 (0.11 - 0.17) |
|  | Two Plus | 3,647 | 0.17 (0.10 - 0.27) | 0.16 (0.10 - 0.23) | 0.20 (0.14 - 0.28) | 0.17 (0.11 - 0.25) |
| Over 65 | All | 66,567 | 0.12 (0.11 - 0.14) | 0.16 (0.14 - 0.18) | 0.17 (0.14 - 0.19) | 0.14 (0.12 - 0.16) |
|  | First | 34,455 | 0.11 (0.08 - 0.13) | 0.12 (0.10 - 0.15) | 0.15 (0.12 - 0.18) | 0.10 (0.07 - 0.13) |
|  | Two Plus | 17,288 | 0.15 (0.12 - 0.19) | 0.20 (0.17 - 0.23) | 0.21 (0.18 - 0.24) | 0.20 (0.15 - 0.26) |

## Conclusions

We believe this is the first example of the escalation antibiogram being applied to community data from the UK and could lead to improved antibiotic prescribing for pyelonephritis, which is an important condition.

Antibiotic resistance and co-resistance patterns will differ from region to region and country to country hence the results of this study are unlikely to be directly transferable. The methodology, however, is simple to apply where the underlying data are routinely collected. If antibiotic resistance among urinary pathogens increases over time, we are likely to see more pyelonephritis due to initial treatment failure for cystitis and to see higher resistance rates to second-line agents. This will increase the utility of our approach to optimise the escalation of antibiotic treatment. We recognise that antibiotic selection decisions are complex, balancing the likely resistance rates against side effect profiles, drug-drug interactions and other patient-specific factors, so these estimated resistance rates will feed into that process, rather than provide a definitive answer to the prescriber.

Within our region we have demonstrated clinically significant differences in rates of expected resistance to pyelonephritis antibiotics when resistance to first-line cystitis antibiotics is accounted for. We suggest this model could be applied to local data in other regions by infection specialists and then simplified to generate practical guidelines for GP, emergency departments and acute medical units.

Within our region we would suggest:

- In pregnancy cefalexin would be first choice and gentamicin second in all cases.
- With suspected or confirmed resistance to pivmecillinam, in any age group, then treat with cefalexin in the first instance and ciprofloxacin second. Avoid the use of co-amoxiclav.
- With suspected or confirmed resistance to trimethoprim then treatment with cefalexin is optimal in patients over 65. In patients under 65, ciprofloxacin has the lowest resistance rate, but either cefalexin or co-amoxiclav may be used due to similar rates of resistance and preferred side effect profiles.
- With suspected or confirmed resistance to nitrofurantoin then use co-amoxiclav first line, with, cefalexin second line. Ciprofloxacin would be a third option.

## Data Availability

All data produced in the present study are available upon reasonable request to the authors

## Ethics

This was a retrospective study using anonymised data already collected as part of routine clinical care. This project was approved by Public Health England’s Research Ethics and Governance Group (REGG).

## Transparency declarations

All authors confirm that they have no conflicts of interest to declare.

## Author Contributions

Conceived the Study and Obtained Funding: P.W., A.W.D., M.B.A.

Cleaning and Analysis of Data: R.B., L.G., A.W.D., P.W.

Drafting of Manuscript: P.W., M.B.A., E.B.

Corrected and Approved Manuscript: All authors

